# Equally low blood metal ion levels at 10-years follow up of total hip arthroplasties with Oxinium, CoCrMo and stainless steel femoral heads. Data from a randomized clinical trial

**DOI:** 10.1101/2022.04.04.22273159

**Authors:** Paul Johan Høl, Geir Hallan, Ove Furnes, Anne Marie Fenstad, Kari Indrekvam, Thomas Kadar

## Abstract

**Background and purpose:** The use of inert head materials such as ceramic heads has been proposed as a method of reducing wear and corrosion products from the articulating surfaces in total hip arthroplasty (THA), as well as from the stem-head taper connection. In the present study, we wanted to evaluate the blood metal ion levels of Oxinium modular femoral heads at 10 years postoperatively and compare with the CoCrMo modular femoral head counterpart, and the monoblock stainless steel Charnley prosthesis.

**Patients and methods:** 150 patients with osteoarthritis of the hip joint previously included in a randomized clinical trial that had the primary aim of examining polyethylene wear in cemented THAs were re-grouped according to femoral head material. One group (n=30) had received the Charnley monoblock stainless steel stem with 22.2 mm head (DePuy, UK). The other groups (n=120) received a Spectron EF CoCrMo stem with either a 28 mm CoCrMo or Oxinium modular head (Smith & Nephew, USA). After 10 years, 19 patients had died and 38 withdraw from the study. Five patients were revised due to infection in their hip and 7 were revised due to aseptic loosening of either the cup or stem or both. 81 patients with median age of 79 years (70-91) were available for whole blood metal ion analysis.

**Results:** The levels of Co, Cr, Ni and Zr in the blood were generally low with all head materials (medians <0.3 micrograms/L). We found no statistically significant difference in the concentration of metal ions between the group with Charnley prosthesis (n=17) and those with Spectron EF stems with either CoCrMo (n=32) or Oxinium heads (n=32) (p=0.18-0.81).

**Interpretation:** In this study of patients with non-revised THAs, no indication of severe trunnion corrosion was found. The blood ion levels with the use of ceramic Oxinium modular femoral heads were similar to CoCrMo or stainless steel heads at 10 years follow-up.

## Introduction

The occurrence of wear and corrosion at the trunnion-head interface (trunnionosis) has gained wider recognition as an important clinical matter in metal-on-metal (Haddad et al. 2011, Dahlstrand et al. 2017, Salib et al. 2018) as well as metal-on-polyethylene THA (Cooper et al. 2012, Weiser et al. 2017). Some suggest that trunnionosis accounts for up to 3 % of all THA failures (Berstock et al. 2018, Sultan et al. 2018). Apart from aseptic loosening and osteolysis from local release of metal debris, also systemic toxicity related to high cobalt ion levels with neurological (Gessner et al. 2019) and cardiac (Lassalle et al. 2018, Choi et al. 2019) manifestations (termed cobaltism), has caused further concerns.

Oxidized zirconium (Oxinium™) femoral heads were introduced with the prospects of combining the strength of a metal head with the smoothness and hardness of ceramic (Jonsson et al. 2015). Oxinium has a metal alloy core (zirconium-niobium) and an approximately 5 µm thick zirconia ceramic surface made by heat treatment of the surface (Good et al. 2003). Theoretically, the Oxinium femoral head and taper junction can reduce the release of metals due to the ceramic surface. One study showed that the use of Oxinium femoral head decreased the blood cobalt levels versus a cobalt-chromium-molybdenum alloy (CoCrMo) femoral head, albeit it did not reach statistical significance (Nam et al. 2015). However, a retrieval study showed no difference in the corrosion score between Oxinium and CoCrMo femoral heads (Tan et al. 2016).

In the present study, we wanted to evaluate the blood metal ion levels of Oxinium modular femoral heads at 10 years postoperatively and compare this with patients with the CoCrMo modular femoral head counterpart, and the monoblock stainless steel Charnley prosthesis. The null hypothesis was that blood metal ion levels with Oxinium femoral heads were equal to that of the metal heads.

## Patients and Methods

The study was part of a randomized study investigating polyethylene wear of five different articulations and migration of the implants. The patient demographics were described in detail in these two publications (Kadar et al. 2011, Kadar et al. 2011).

The trial was registered with ClinicalTrials.gov (NCT00698672) and approved by the Regional Comittee for Medical Research Ethics Western Norway. An amendment to the protocol, which included blood metal analysis, MRI and CT at 10-year follow-up was also approved (REK number 2014-02370). Between November 2004 and June 2007, 150 patients with primary or secondary osteoarthritis of the hip were included in the study (Figure 1). Each patient provided informed consent to participation in the study. Exclusion criteria were BMI >35, uncompensated cardio-pulmonary disease, malignant disease, dementia, rheumatoid arthritis or other serious systemic diseases.

**Figure 1.**
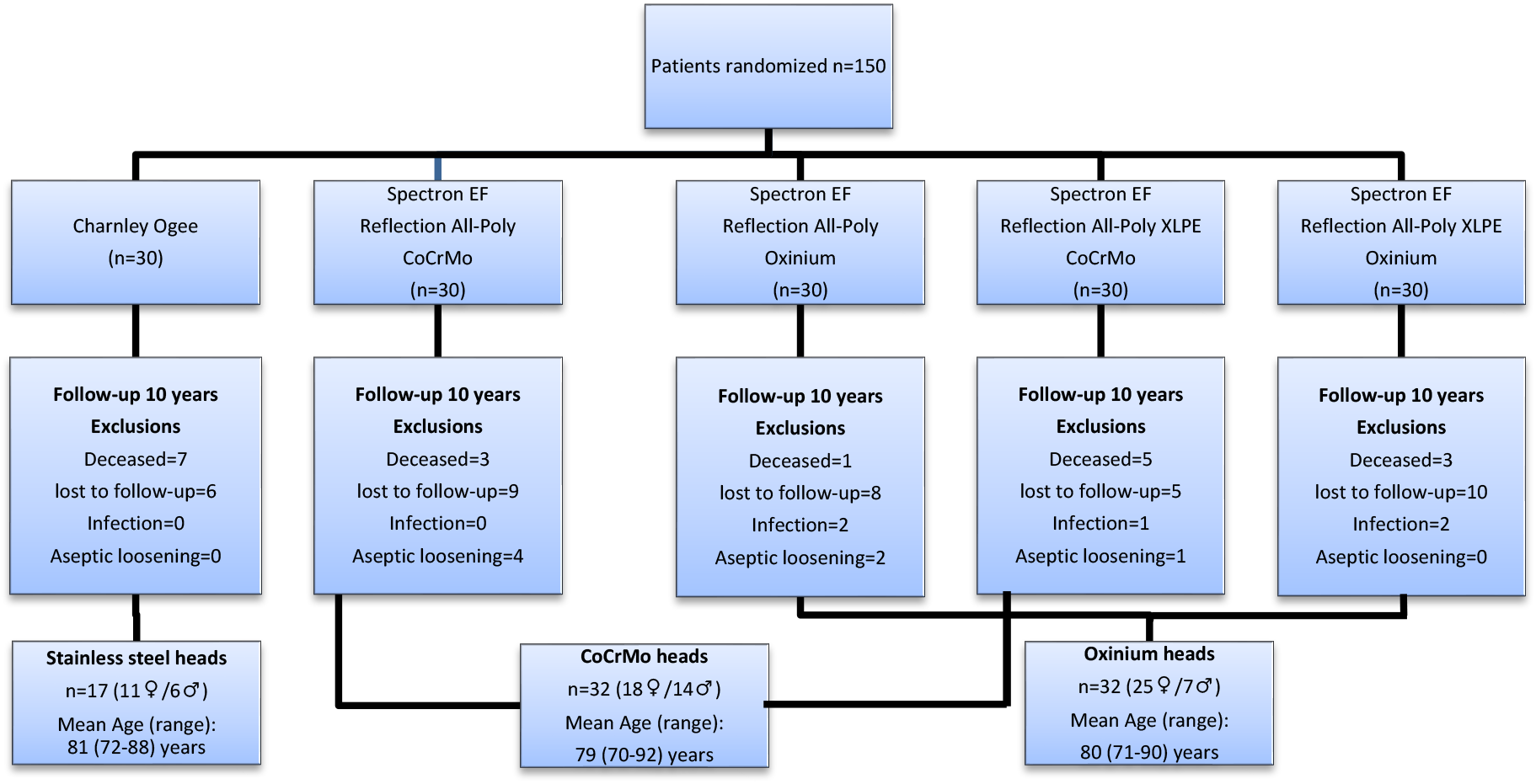
Participant flow diagram describing the original five study groups based on the different head-cup articulations and the final three groups based on head material (stainless steel, CoCrMo and Oxinium). Exclusions after 10 years follow up are sorted after revision causes (infection or aseptic loosening) and reasons for drop out (deceased or lost to follow-up). The gender distribution and age (mean and range) in the three study groups are specified.

The patients were randomized into five groups of cemented THAs with n=30 in each group. One group received the Charnley flanged 40 femoral 316L stainless steel monoblock stem with a 22.2 mm head, which articulated with a Charnley Ogee UHMWPE acetabular cup (DePuy, UK) (Figure 2a). The other four groups received a Spectron EF CoCrMo femoral stem (Figure 2b) with either a 28 mm CoCrMo or a 28 mm Oxinium femoral head (Figure 2c), which articulated with either an ethylene oxide-sterilized Reflection All-Poly UHMWPE cup or a Reflection All-Poly highly cross-linked polyethylene (XLPE) cup (Smith&Nephew, UK) (Figure 2 d).

**Figure 2.**
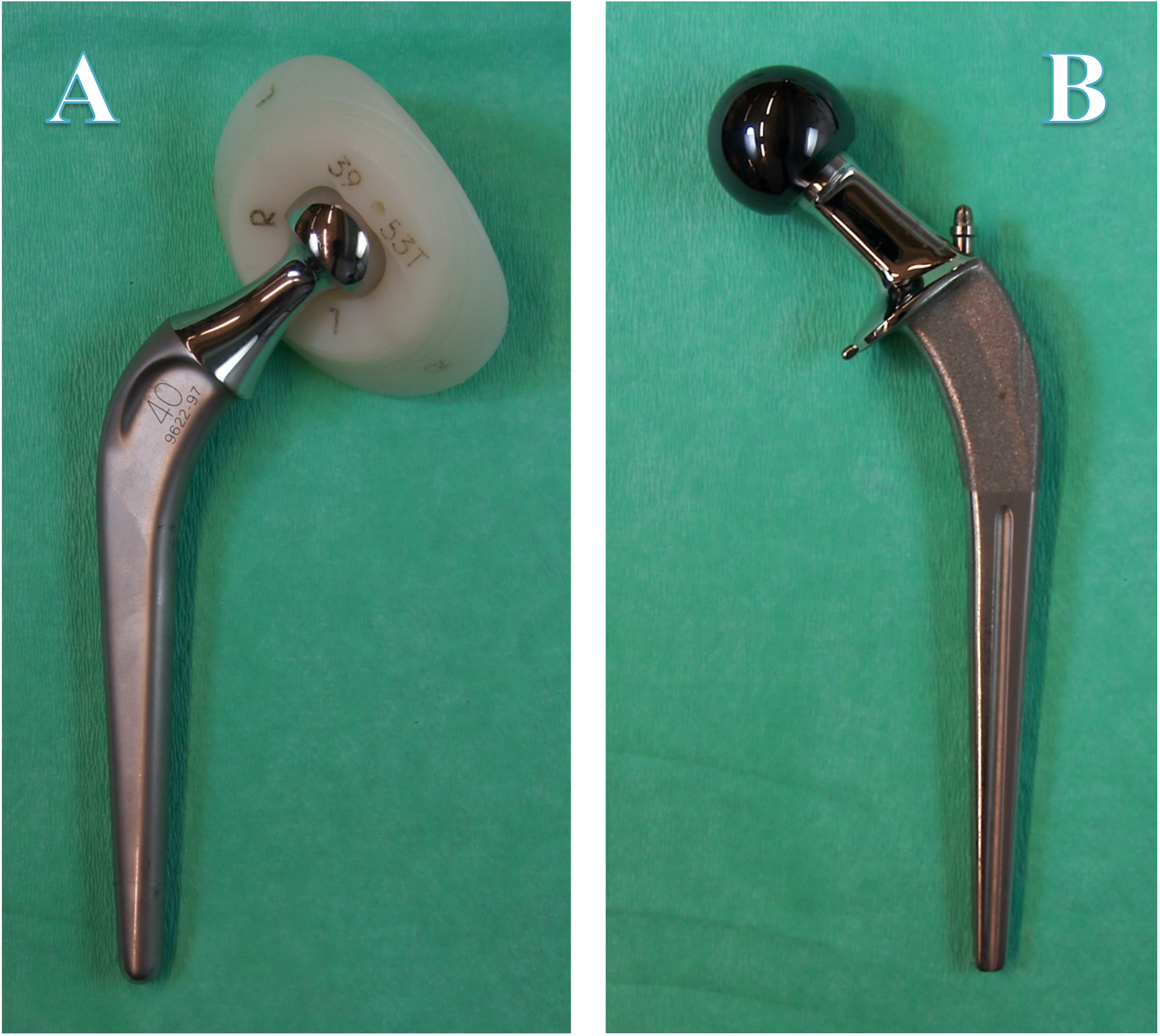
A. Charnley 40 femoral 316L stainless steel monoblock stem with a 22.2 mm head, which articulated with a Charnley Ogee UHMWPE acetabular cup. B. Spectron EF CoCrMo femoral stem with a 28 mm Oxinium modular femoral head

**Figure 2.**
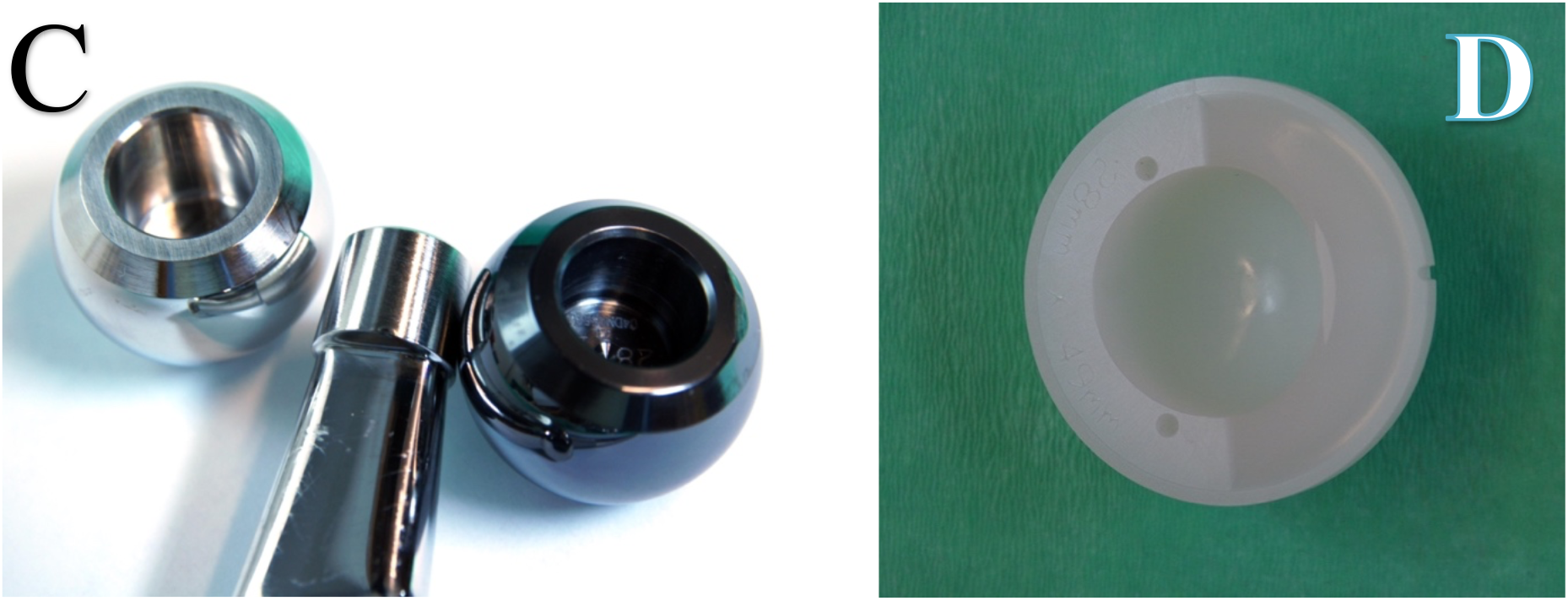
C. CoCrMo 28 mm femoral head (left) and Oxinium 28 mm femoral head (right). Spectron EF stem taper (middle). D. Reflection All-Poly UHMWPE cup

Only patients suitable to receive both the Spectron EF stem size 2–5 with a standard offset and the Charnley Flanged 40 stem were recruited in the study. The surgical procedure, post-operative treatment and method of wear measurement has previously been described in detail (Kadar 2011).

Of the 150 patients in the original study, 19 patients had deceased and 38 patients withdraw from the study (not able or willing to participate) (Figure 1). Seven patients were revised before 10 years follow up due to aseptic loosening of either the cup or stem or both. Four of these cases were included in a retrieval project (Wolf et al. 2022). Additionally, five hips were revised due to infection. None of the patients in the Charnley group were revised.

After 10-year follow-up, the remaining 81 patients (54 females (67%) and 27 men (33%) from the original five groups were regrouped into three groups, depending on the femoral head material used at the time of surgery (Figure 1). The overall median age were 79 years (70-91). At 10 years follow-up we detected that many patients had received other implants, from data at the Norwegian Arthroplasty Register (supplement 1).

### Blood metal ion analysis

Whole blood was drawn from a vein in the forearm using an intravenous catheter (venesection) in our hospital (Becton Dickinson Venflon Pro™, 1.5 × 45 mm, Helsingborg, Sweden). After venipuncture, the steel needle was removed, and the blood (5-10 mL) was allowed to flow directly into a 15 ml metal free polypropylene tube (VWR International, CS500, Radnor, PA, US). The blood was stored at minus 20ºC until further handling. The concentration of chromium, cobalt, nickel and zirconium in whole blood was determined by Inductively Coupled Plasma - Mass Spectrometry (ICP-MS). The instrument (Element XR™ HR-ICP-MS, Thermo Fisher Scientific, Bremen, Germany) was equipped with a high resolution, magnetic sector field. Blood metal concentrations are available upon request. Before analysis, the blood was acid digested in closed vessels in a microwave-assisted system (Milestone 1200 Mega, Sorisole, Italy). An aliquot of approx. 1.5 g whole blood was added 3 mL 60% ultrapure HNO_3_ and 2 mL 30% H_2_O_2_ (Merck, Darmstadt, Germany). After digestion the samples were diluted approx. 66 times with double distilled deionized water (MilliQ, USA). Calibration was performed by standard addition using 0.2, 0.5, 2 and 5 µg/L calibration solutions prepared from a 10 µg/mL multi-element stock standard solution (Spectrascan, Lot Number C2-MEB294066, Teknolab AB, Kungsbacka, Sweden). An internal standard of indium was injected automatically to all the samples to monitor and correct for any instrumental changes. The method detection limit (MDL) for element analysis was defined as three times the standard deviation in ten blank solutions measured at different times, taking the dilution into account. The MDL was 0.11 µg/L for Cr, 0.04 µg/L for Co, 0.21 for Ni and 0.02 for Zr. The accuracy of the analytical method was monitored using a reference material (Seronorm Trace Elements Whole Blood L-2, lot 1103129, Sero AS, Billingstad, Norway).

### Statistics

All statistical analyses were performed using the IBM SPSS Statistics for Macintosh, Version 27.0 (IBM Corp, Armonk, NY, USA). The graph was made with GraphPad Prism software, version 9.3.1 (GraphPad Software Inc., San Diego, CA, USA). Since the metal concentrations in the study groups had a skewed distribution and did not pass the normality test, nonparametric statistical methods were chosen. Kruskal–Wallis one-way analysis of variance was used to detect differences in average blood ion levels among the three study groups. Additionally, the groups with different cup materials were also compared with Kruskal-Wallis test. An independent samples Mann-Whitney U test was used to compare metal ion levels between patients with prostheses in other joints (pooled) versus patients with the study prosthesis only (pooled). Probability (P) values of <0.05 denote statistical significance.

## Results

We found no statistical differences in blood ion levels between the stainless steel Charnley prosthesis and the Spectron EF stems with either CoCrMo or Oxinium femoral head (p=0.18-0.81) (Table 1 and Figure 3). The blood metal ion levels for Cr, Co, Zr and Ni were generally low in all study groups (all medians <0.3 microgram/L). The different acetabular cup materials (conventional or highly crosslinked polyethylene) did not influence the metal levels (p=0.18-0.89, five different articulations).

**Table 1.** Co, Cr, Zr and Ni concentrations in the 3 groups based on head material (stainless steel, CoCrMo and Oxinium), represented with mean, median and maximum values (microgram/L). Statistical difference between the groups is calculated by Kruskal-Wallis one-way test.

|  | **Cr** | **Co** | **Zr** | **Ni** |
| --- | --- | --- | --- | --- |
| **Stainless steel heads (n=17)** |  |  |  |  |
| Median | 0.14 | 0.08 | 0.05 | 0.24 |
| Mean | 0.38 | 0.25 | 0.32 | 0.29 |
| max | 1.02 | 1.96 | 2.30 | 0.62 |
| **CoCrMo heads (n=32)** |  |  |  |  |
| Median | 0.29 | 0.15 | 0.12 | 0.17 |
| Mean | 0.39 | 0.39 | 0.36 | 0.31 |
| max | 1.52 | 2.07 | 2.19 | 2.31 |
| **Oxinium heads (n=32)** |  |  |  |  |
| Median | 0.17 | 0.13 | 0.20 | 0.20 |
| Mean | 0.33 | 0.28 | 0.41 | 0.26 |
| max | 1.95 | 1.31 | 3.68 | 1.14 |
| **P-values (Kruskal-Wallis)** | 0.44 | 0.26 | 0.18 | 0.81 |

**Figure 3.**
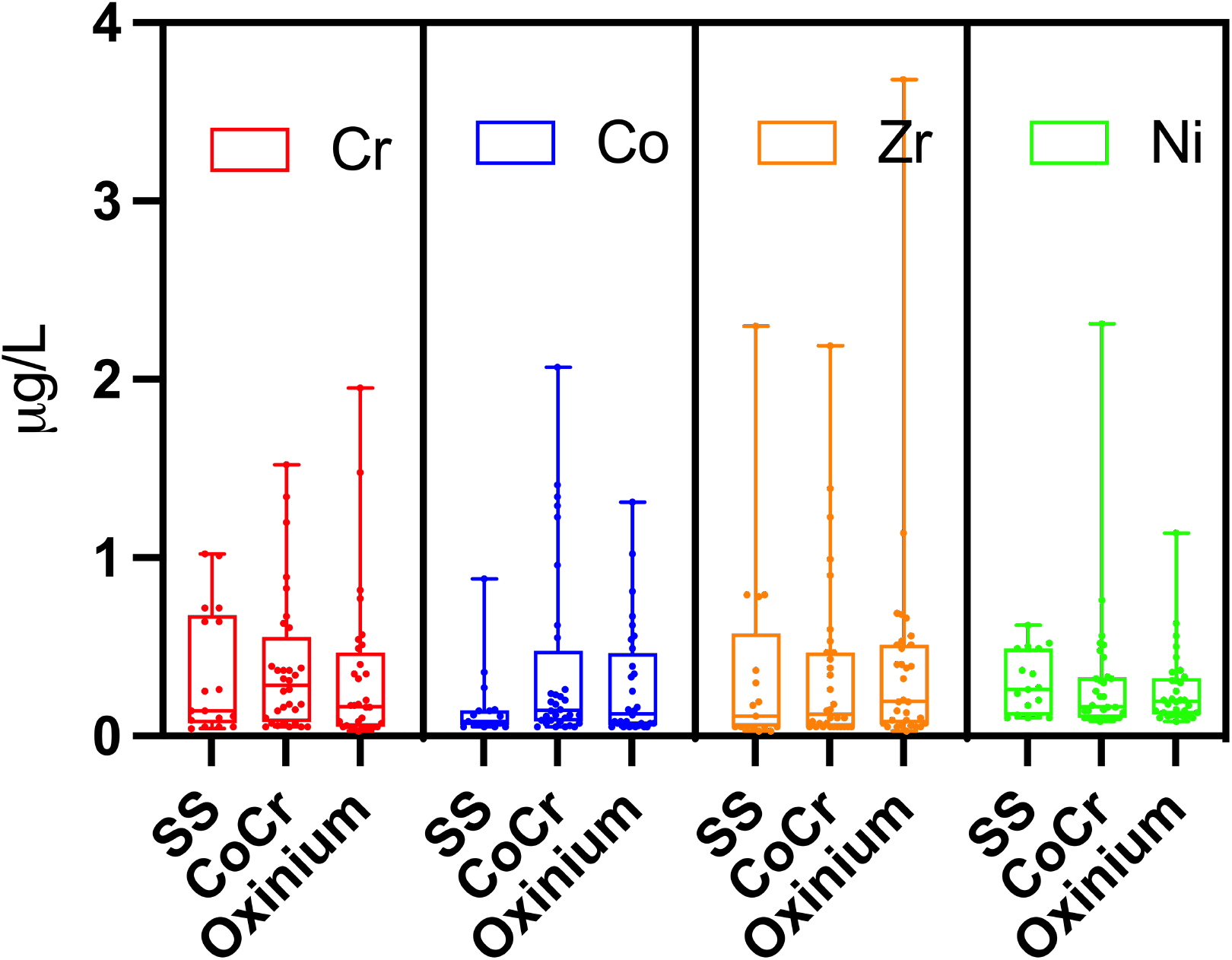
Boxplot of blood metal ion levels (ug/L) for Cr, Co, Zr and Ni, grouped on head material. Horizontal lines inside the box indicates median level. Whiskers represents min-max values (range).

Many study patients had prostheses in other joints, surgery performed either before or within the study period (55 of 81, 67.9%), see Supplement 1 for further details. 50 patients had a contralateral hip replacement. The femoral stems used in the other hip was the Lubinus SP II (n=23), Spectron EF (n=13), Charnley (n=6), Exeter (n=3), Profile (n=3), Titan, Corail, Elite and MS-30 (n=1). Eleven patients had a total knee replacement; Profix (n=5), NexGen (n=3), LCS Complete (n=3) and Genesis (n=2). Nine of these had bilateral hip replacement. Three patients also had shoulder prosthesis. The patient group with Oxinium femoral heads had the highest percentage of prostheses in other joints (23/32=71.9%), followed by the Charnley stainless steel group (12/17=70.6%) and the CoCrMo group (20/32=62.5%).

In patients who had additional prostheses (n=55) the median metal ion levels were not statistically different (p=0.15-0.85) to those with only the study prosthesis (n=26) (Supplement 2).

## Discussion

In the present study of non-revised primary THAs we found no difference in blood ion levels with the use of Oxinium femoral heads compared to CoCrMo or stainless steel heads at 10 years follow-up. All the Co and Cr values were below well below the British Medicines and Healthcare Products Regulatory Agency threshold value of 7 μg/L, and all the median values for the 3 head materials were similar and all below 0.3 μg/L.

The mechanisms of trunnionosis are currently poorly understood and is likely multi-factorial (Mistry et al. 2016). Several Electrochemical and mechanical factors have been suggested to increase the risk. Implant-based factors that have been postulated to affect the degree of trunnionosis include length and diameter of the head, the dimensions of the trunnion, the surface finish and flexural rigidity of the trunnion and the head-trunnion interface materials (Lanting et al. 2017, Weiser and Lavernia 2017). Patients with CoCr-containing THAs had more necrosis and lymphocytic infiltration in their tissues than patients with hip resurfacings, indicating that trunnion wear debris produced more cytotoxic and immunogenic reactions than bearing wear debris (Lehtovirta et al. 2018). Especially the cobalt species has been under suspicion for its adverse effects, such as cardiac toxicity (Lassalle 2018). However, recent data from the National Joint Registry and NHS English hospital inpatient episodes about CoCr-containing THAs, did not find increased risk of all-cause mortality, or clinically meaningful heart outcomes, cancer, or neurodegenerative disorders into the second decade post-implantation (Deere et al. 2022).

One study has suggested that by using a ceramic femoral head, CoCrMo tribocorrosion may be mitigated (Kurtz et al. 2013). Two other studies also concluded that ceramic head confers an advantage in trunnion fretting and corrosion (Tan 2016, Morlock et al. 2020).

Mid-term *in vivo* wear results (2- and 5-year follow-up based on radiostereometric analysis (RSA)) have not shown wear benefits of Oxinium femoral heads over conventional CoCrMo femoral heads (Kadar 2011, Jonsson 2015). There are reports that Oxinium femoral heads are susceptible to damage upon hip dislocation with subsequent increased polyethylene wear (Gibon et al. 2013). A recent case report showed catastrophic failure of an Oxinium-coated total knee prosthesis (Smith and Nephew) that resulted in metallosis with extra-articular extravasation (Purcell et al. 2020). In a systematic review and meta-analysis, Oxinium femoral heads did not lead to lower polyethylene wear rate or higher survival rate, when compared with CoCr femoral heads in patients treated with THA (Malahias et al. 2019). The evidence of any benefits with the use of Oxinium femoral heads is therefore still sparse and further investigation regarding any benefit with Oxinium femoral heads is therefore needed. That said, clinical data from the Australian and UK registry indicate excellent results for Oxinium used in combination with well-performing cup and stems (Australian Orthopaedic Association National Joint Replacement Registry 2019, Davis et al. 2020).

Furthermore, it is now well established that highly cross-linked polyethylene (HXLPE) acetabular cups and liners significantly reduces wear compared to conventional polyethylene (CPE). (McCalden 2018) This has allowed for the use of larger head sizes to improve prosthetic stability. However, larger femoral head sizes may increase stress at the modular head–neck junction, increasing the effective horizontal lever arm, potentially leading to fretting corrosion and metal debris with the use of cobalt-alloy femoral heads (Dyrkacz et al. 2013, Morlock 2020). In this study only small head sizes were used. Thus, we cannot deduct our results to larger head sizes. However, in a recent study studying metal ion levels with modular 28 mm, 36 mm and 40 mm CoCrMo femoral heads, no difference in cobalt and chromium levels were found with increasing femoral head sizes after 10 years follow-up (Dover et al. 2020).

Neck taper flexural rigidity also affect trunnion corrosion and one study showed that lower stem flexural rigidity predicted stem fretting and corrosion damage in a ceramic head cohort, but not in the metal head cohort (Kurtz 2013). Care should therefore be taken to generalize the results to different stem designs and materials.

### Strengths and limitations

We did not have a control group of patients with native hip, without implants. Yet, the monoblock stainless steel Charnley prosthesis had the same metal ion levels as the modular Spectron EF prosthesis, regardless of femoral head material. Trunnionosis cannot account for the metal ion levels with the monoblock Charnley, showing that systemic metal ions may stem from other sources than the trunnion. The Spectron EF femoral stem was more stable than the Charnley stem in a RSA-study when evaluated at 2 years (Kadar 2011). Theoretically the blood ion levels with the Charnley prosthesis may be the result of corrosion at the stem-cement interface. Analysis of the retrieved prostheses has been undertaken to clarify this matter in another study. Cases with Spectron EF stem loosening had significantly higher median Cr (1.05 μg/L), Co (1.85 μg/L) and Zr (0.65 μg/L) concentrations in blood at the day of revision surgery compared to the cases with the same femoral stem in this study (Wolf 2022). It was concluded that fretting wear of loose stems against the cement mantle may explain the elevated metal ion levels.

An interesting research question for a future study of our study patients is to investigate if there is an association between implant migration measured by radiostereometry, and blood metal ion levels.

Another limitation with the study is that 50 patients had hip prosthesis in the other hip. Some patients also had prostheses in the knee (n=11) and in the shoulder (n=3), see Supplement 1 for details about implant brands. This can contribute extra to the total metal burden in the blood and might hide differences between the study groups, although this doesn’t seem to be the case, since individuals who only had the study prosthesis did not have significantly lower metal levels (supplement 2). Despite potential for metal contamination from other implants, the metal levels were low in all groups, ruling out there is a serious problem with metal ion release due to trunnionosis with our study prostheses at 10 years post-op.

One strength of this study is that it was part of a randomized study, reducing the risk of selection bias. Another strong point of this study is that the taper design was identical between the CoCrMo and Oxinium modular head groups. Taper design is associated with the degree of crevice corrosion at the modular head and neck interface. Our results are therefore solely associated with head-neck material, not variance in taper design.

Detecting differences in blood ion levels between groups is demanding due to small detection limits (Michel et al. 1991). In the present study a single laboratory, with a highly sensitive methodology and adequate detection limits, analyzed all blood samples. This allows more subtle differences between groups to be detected.

Our study patients were quite old, so our results may not apply to younger and more active patients, who are increasingly becoming eligible for the THA procedure. The mechanical demands at the head-trunnion in a younger population interface may be higher. Thus, any mechanically driven corrosion and rise in blood ion levels may be underestimated in the population of this study.

In this study of well-functioning THAs with small femoral heads, no indication of severe trunnion corrosion was found. Although we found no difference in blood ion levels with the use of modular Oxinium or metal femoral heads, further investigation of periprosthetic tissues and retrieved implants may give insight to any differences in wear and corrosion mechanism at the trunnion-head interface between CoCrMo and Oxinium femoral heads. Our results highlight the importance of retrieval centers for understanding cause and effect relationships in THA (Ellison et al. 2012, Wolf 2022).

## Data Availability

All data produced in the present study are available upon reasonable request to the authors.

## Author contributions

PJH: Collected and interpreted the data, Performed the statistical analysis, created the figures, carried out literature search, wrote the paper.

TK: Designed the study, provided clinical review of patients in clinic, wrote the paper, carried out literature search.

GH, OF and KI: Designed the study, revised the manuscript

AMF: Provided patient details from initial study, controlled the statistical analysis.

## Acknowledgments

We would like to thank the surgeons at Bergen Hospital Trust (Helse Bergen HF) and the subject participants for their support in conducting this research. The study was supported financially by Western Norway Regional Health Authority, Smith & Nephew Norway and OrtoMedic AS. None of the sources of funding were involved in the planning or analysis of results. Terje Stokke (Radiology dept., HUS) were responsible for taking the blood samples and Irene Ohlen Moldestad performed the blood sample preparation. The authors also thank senior engineer Siv Hjorth Dundas for her excellent technical support related to the ICP-MS analysis at the Department of Earth Sciences, UiB.

## Data Availability Statement

The data that support the findings of this study are available upon request from the corresponding author.

