## Supplement 1 and 2 for "Equally low blood metal ion levels at 10-years follow up of total hip arthroplasties with Oxinium, CoCrMo and stainless steel femoral heads. Data from a randomized clinical trial"

**Supplement 1.** Overview of included study patients (n=81) with their study prosthesis (femoral stem, head and cup) grouped by femoral head material. Further columns list additional hip stems (SS=stainless steel), knee and shoulder prostheses implanted either before or within the 10-year study period.

| Patient | Stem | Head material | Cup | 2nd hip stem (stem material) | 1st knee | 2nd knee | Shoulder |
|---|---|---|---|---|---|---|---|
| 1 | Charnley flanged 40 | **Stainless steel, 316L** | Charnley Ogee UHMWPE | LUBINUS SP II (CoCrMo) | | | |
| 2 | Charnley flanged 40 | **Stainless steel, 316L** | Charnley Ogee UHMWPE | PROFILE (TiAlV) | Genesis I (CoCr/TiAlV) | | |
| 3 | Charnley flanged 40 | **Stainless steel, 316L** | Charnley Ogee UHMWPE | | | | |
| 4 | Charnley flanged 40 | **Stainless steel, 316L** | Charnley Ogee UHMWPE | | | | |
| 5 | Charnley flanged 40 | **Stainless steel, 316L** | Charnley Ogee UHMWPE | CHARNLEY (SS) | PROFIX (CoCrMo) | PROFIX (CoCrMo) | |
| 6 | Charnley flanged 40 | **Stainless steel, 316L** | Charnley Ogee UHMWPE | LUBINUS SP II (CoCrMo) | | | |
| 7 | Charnley flanged 40 | **Stainless steel, 316L** | Charnley Ogee UHMWPE | | PROFIX (CoCrMo) | | |
| 8 | Charnley flanged 40 | **Stainless steel, 316L** | Charnley Ogee UHMWPE | LUBINUS SP II (CoCrMo) | | | |
| 9 | Charnley flanged 40 | **Stainless steel, 316L** | Charnley Ogee UHMWPE | LUBINUS SP II (CoCrMo) | | | |
| 10 | Charnley flanged 40 | **Stainless steel, 316L** | Charnley Ogee UHMWPE | LUBINUS SP II (CoCrMo) | | | |
| 11 | Charnley flanged 40 | **Stainless steel, 316L** | Charnley Ogee UHMWPE | | LCS (CoCrMo) | | |
| 12 | Charnley flanged 40 | **Stainless steel, 316L** | Charnley Ogee UHMWPE | | | | |
| 13 | Charnley flanged 40 | **Stainless steel, 316L** | Charnley Ogee UHMWPE | | | | |
| 14 | Charnley flanged 40 | **Stainless steel, 316L** | Charnley Ogee UHMWPE | ELITE (CoCrMo) | | | |
| 15 | Charnley flanged 40 | **Stainless steel, 316L** | Charnley Ogee UHMWPE | LUBINUS SP II (CoCrMo) | | | |
| 16 | Charnley flanged 40 | **Stainless steel, 316L** | Charnley Ogee UHMWPE | LUBINUS SP II (CoCrMo) | | | |
| 17 | Charnley flanged 40 | **Stainless steel, 316L** | Charnley Ogee UHMWPE | | | | |
| 18 | Spectron EF | **CoCrMoMo** | Reflection All-Poly UHMWPE | | | | |
| 19 | Spectron EF | **CoCrMoMo** | Reflection All-Poly UHMWPE | LUBINUS SP II (CrCo) | LCS (CoCrMo) | | |
| 20 | Spectron EF | **CoCrMoMo** | Reflection All-Poly UHMWPE | | | | |
| 21 | Spectron EF | **CoCrMoMo** | Reflection All-Poly UHMWPE | LUBINUS SP II (CoCrMo) | PROFIX (CoCrMo) | PROFIX (CoCrMo) | |
| 22 | Spectron EF | **CoCrMoMo** | Reflection All-Poly UHMWPE | | | | |
| 23 | Spectron EF | **CoCrMoMo** | Reflection All-Poly UHMWPE | LUBINUS SP II (CoCrMo) | | | |
| 24 | Spectron EF | **CoCrMoMo** | Reflection All-Poly UHMWPE | LUBINUS SP II (CoCrMo) | | | |
| 25 | Spectron EF | **CoCrMoMo** | Reflection All-Poly UHMWPE | SPECTRON-EF (CoCrMo) | | | |
| 26 | Spectron EF | **CoCrMoMo** | Reflection All-Poly UHMWPE | SPECTRON-EF (CoCrMo) | | | |
| 27 | Spectron EF | **CoCrMoMo** | Reflection All-Poly UHMWPE | | | | |
| 28 | Spectron EF | **CoCrMoMo** | Reflection All-Poly UHMWPE | LUBINUS SP II (CoCrMo) | | | |
| 29 | Spectron EF | **CoCrMoMo** | Reflection All-Poly UHMWPE | | | | |
| 30 | Spectron EF | **CoCrMoMo** | Reflection All-Poly UHMWPE | LUBINUS SP II (CoCrMo) | | | |
| 31 | Spectron EF | **CoCrMoMo** | Reflection XLPE | | | | |
| 32 | Spectron EF | **CoCrMoMo** | Reflection XLPE | LUBINUS SP II (CoCrMo) | | | |
| 33 | Spectron EF | **CoCrMoMo** | Reflection XLPE | | | | Global Adv. (CoCrMo) |
| 34 | Spectron EF | **CoCrMoMo** | Reflection XLPE | SPECTRON-EF (CoCrMo) | | | |
| 35 | Spectron EF | **CoCrMoMo** | Reflection XLPE | CHARNLEY (SS) | | | |
| 36 | Spectron EF | **CoCrMoMo** | Reflection XLPE | | | | |
| 37 | Spectron EF | **CoCrMoMo** | Reflection XLPE | SPECTRON-EF (CoCrMo) | | | |
| 38 | Spectron EF | **CoCrMoMo** | Reflection XLPE | | | | |
| 39 | Spectron EF | **CoCrMoMo** | Reflection XLPE | LUBINUS SP II (CoCrMo) | | | |
| 40 | Spectron EF | **CoCrMoMo** | Reflection XLPE | LUBINUS SP II (CrCo) | | | |
| 41 | Spectron EF | **CoCrMoMo** | Reflection XLPE | | | | |
| 42 | Spectron EF | **CoCrMoMo** | Reflection XLPE | CHARNLEY (SS) | | | |
| 43 | Spectron EF | **CoCrMoMo** | Reflection XLPE | | | | |
| 44 | Spectron EF | **CoCrMoMo** | Reflection XLPE | | | | |
| 45 | Spectron EF | **CoCrMoMo** | Reflection XLPE | SPECTRON-EF (CoCrMo) | | | |
| 46 | Spectron EF | **CoCrMoMo** | Reflection XLPE | | | | |
| 47 | Spectron EF | **CoCrMoMo** | Reflection XLPE | EXETER (SS) | | | |
| 48 | Spectron EF | **CoCrMoMo** | Reflection XLPE | LUBINUS SP II (CoCrMo) | NexGen (TiAlV) | NexGen (TiAlV) | |
| 49 | Spectron EF | **CoCrMoMo** | Reflection XLPE | MS-30 (SS) | | | |
| 50 | Spectron EF | **Oxinium** | Reflection All-Poly UHMWPE | | | | |
| 51 | Spectron EF | **Oxinium** | Reflection All-Poly UHMWPE | SPECTRON-EF (CoCrMo) | Genesis I (CoCr/TiAlV) | | |
| 52 | Spectron EF | **Oxinium** | Reflection All-Poly UHMWPE | PROFILE (TiAlV) | | | |
| 53 | Spectron EF | **Oxinium** | Reflection All-Poly UHMWPE | Profile/Tri-lock plus (TiAlV) | | | |
| 54 | Spectron EF | **Oxinium** | Reflection All-Poly UHMWPE | TITAN (TiAlV) | | | |
| 55 | Spectron EF | **Oxinium** | Reflection All-Poly UHMWPE | SPECTRON-EF (CoCrMo) | | | |
| 56 | Spectron EF | **Oxinium** | Reflection All-Poly UHMWPE | | | | |
| 57 | Spectron EF | **Oxinium** | Reflection All-Poly UHMWPE | EXETER (SS) | | | Global Adv. (CoCrMo) |
| 58 | Spectron EF | **Oxinium** | Reflection All-Poly UHMWPE | CHARNLEY (SS) | | | |
| 59 | Spectron EF | **Oxinium** | Reflection All-Poly UHMWPE | | | | |
| 60 | Spectron EF | **Oxinium** | Reflection All-Poly UHMWPE | LUBINUS SP II (CoCrMo) | | | |
| 61 | Spectron EF | **Oxinium** | Reflection All-Poly UHMWPE | | | | |
| 62 | Spectron EF | **Oxinium** | Reflection All-Poly UHMWPE | SPECTRON-EF (CoCrMo) | | | |
| 63 | Spectron EF | **Oxinium** | Reflection All-Poly UHMWPE | | | | Delta Xtend (CoCrMo) |
| 64 | Spectron EF | **Oxinium** | Reflection All-Poly UHMWPE | CORAIL (TiAlV) | PROFIX (CoCrMo) | NexGen (TiAlV) | |
| 65 | Spectron EF | **Oxinium** | Reflection All-Poly UHMWPE | LUBINUS SP II (CoCrMo) | | | |
| 66 | Spectron EF | **Oxinium** | Reflection All-Poly UHMWPE | LUBINUS SP II (CoCrMo) | | | |
| 67 | Spectron EF | **Oxinium** | Reflection XLPE | | | | |
| 68 | Spectron EF | **Oxinium** | Reflection XLPE | EXETER (SS) | NexGen (TiAlV) | | |
| 69 | Spectron EF | **Oxinium** | Reflection XLPE | SPECTRON-EF (CoCrMo) | LCS (CoCrMo) | | |
| 70 | Spectron EF | **Oxinium** | Reflection XLPE | | | | |
| 71 | Spectron EF | **Oxinium** | Reflection XLPE | SPECTRON-EF (CoCrMo) | | | |
| 72 | Spectron EF | **Oxinium** | Reflection XLPE | SPECTRON-EF (CoCrMo) | | | |
| 73 | Spectron EF | **Oxinium** | Reflection XLPE | CHARNLEY (SS) | | | Global Adv. (CoCrMo) |
| 74 | Spectron EF | **Oxinium** | Reflection XLPE | LUBINUS SP II (CoCrMo) | | | |
| 75 | Spectron EF | **Oxinium** | Reflection XLPE | | PROFIX (CoCrMo) | | |
| 76 | Spectron EF | **Oxinium** | Reflection XLPE | | | | |
| 77 | Spectron EF | **Oxinium** | Reflection XLPE | LUBINUS SP II (CoCrMo) | | | |
| 78 | Spectron EF | **Oxinium** | Reflection XLPE | SPECTRON-EF (CoCrMo) | | | |
| 79 | Spectron EF | **Oxinium** | Reflection XLPE | | | | |
| 80 | Spectron EF | **Oxinium** | Reflection XLPE | CHARNLEY (SS) | | | |
| 81 | Spectron EF | **Oxinium** | Reflection XLPE | | | | |

**Supplement 2.** A comparison of the of the blood metal levels (µg/l) in patients with only the study prosthesis (pooled) and those with additional prostheses (pooled). An independent samples Mann-Whitney U test was used to compare ranks.

| | **Groups** | **N** | **Medians (µg/l)** | **Mean ranks** | **p-value** |
|---|---|---|---|---|---|
| Chromium | Only study prosthesis | 26 | 0.17 | 40.27 | 0.85 |
| | Additional prostheses | 55 | 0.20 | 41.35 | |
| Cobalt | Only study prosthesis | 26 | 0.11 | 40.13 | 0.82 |
| | Additional prostheses | 55 | 0.14 | 41.41 | |
| Zirconium | Only study prosthesis | 26 | 0.07 | 35.58 | 0.15 |
| | Additional prostheses | 55 | 0.19 | 43.56 | |
| Nickel | Only study prosthesis | 26 | 0.17 | 38.12 | 0.45 |
| | Additional prostheses | 55 | 0.20 | 42.36 | |
